# Body Mass Index Asian Populations category and stroke and heart disease in the adult population: A longitudinal study of The Indonesia Family Life Survey (IFLS) 2007 and 2014

**DOI:** 10.1101/2023.10.31.23297817

**Authors:** Kamaluddin Latief, Dieta Nurrika, Min-Kuang Tsai, Wayne Gao

## Abstract

**Background:** Stroke and heart disease are included in the Global Burden of Diseases, Injuries, and Risk Factors Study (GBD) concerns. Body Mass Index (BMI) is a modifiable risk factor for stroke and heart disease alike. Most studies classify BMI according to the WHO BMI cut-off point in stroke and heart disease studies. However, there is a limited understanding of the association between the BMI cut-off point in the Asian population category and stroke and heart disease. This study aimed to investigate the incidence rate ratio of stroke and heart disease by BMI categories for Asian population.

**Methods:** A 7-year prospective longitudinal study (2007-2014) was conducted on 6,688 adult Indonesian individuals (≥ 35 years) living in 13 different provinces in Indonesia during the survey periods. Data on BMI were collected in 2007. Information on stroke and heart disease was obtained in both 2007 and in 2014. A multivariate-adjusted Poisson regression model was used to estimate the incidence rate ratio (IRR) and 95% confidence intervals (CIs) of either stroke or heart disease or both stroke and heart disease by BMI.

**Results:** Out of 6,688 eligible participants, 334 (5%) were judged as stroke and heart disease in 2014. The IRR (95% CI) of stroke and heart disease for participants with obesity was 2.57 (1.64-4.04) compared with those within normal weight. This incidence rate ratio remained among middle-aged adults (<55 years) rather than the older adults (≥55 years); the IRR of stroke and heart disease among obese middle-aged adults was 4.18 (95% CI 2.10–8.31).

**Conclusions:** The association was observed between obesity and the risk of stroke and heart disease, especially in middle-aged adults. These findings suggest that lowering BMI through healthy dietary habits and increasing physical activity, especially among middle-aged adults with high education, who are employees, and who live in urban or rural areas may be beneficial for preventing stroke and heart disease.

## Background

Cardiovascular diseases (CVDs) are the leading cause of death globally. An estimated 17.9 million people died from CVDs in 2019, representing 32% of all global deaths. Of these deaths, 85% were due to stroke and heart disease. About 7.4 million deaths, or 3.2% of the total, were due to ischemic heart disease (IHD). In 2019, IHD was recorded as the single highest cause of death, with around 9.0 million deaths [1].

Stroke was the third most frequent cause of disability worldwide and the second most frequent cause of death [2]. The Global Burden of Diseases, Injuries, and Risk Factors Study (GBD) from 1990-2019 found that the absolute number of incidents of stroke increased by 70%, the prevalence of stroke increased by 85%, deaths from stroke increased as well, by 43%, and the cause of death and disability (DALYs) due to stroke increased by 32% [3].

Indonesia is the largest country in South East Asia, with the fourth biggest population in the world [4]. According to the Indonesian Basic Health Research Survey (*Riskesdas*), the prevalence of non-communicable diseases (NCDs) has significantly increased among the Indonesian population between 2007 and 2013, from 8.3% to 12.1% [5, 6], respectively. In Indonesia, the three leading causes of DALYs in 2016 were ischemic heart disease, cerebrovascular disease, and diabetes [7].

Body Mass Index (BMI) is one of the modifiable risk factors for stroke and heart disease [8, 9]. Moreover, previous studies have reported that BMI was associated with stroke and heart disease [10–14]. Furthermore, most previous studies on BMI and stroke and heart disease, conducted in Europe, America, and Asia [14], classify BMI according to the WHO BMI cut-off points. In general, the Asian population has a lower BMI cut-off-point than the non-Asian population [15]. Therefore, WHO experts recommend the BMI cut-off point of Asian populations for public health action [16].

In Indonesia, a previous study using IFLS data found that between 1993 and 2014, the prevalence of overweight people doubled from 17.1% to 33.0%, and the younger generations have higher BMIs than the older generations [17]. The mean BMI also increased from 21.4 kg/m^2^ in 1993 to 23.85 kg/m^2^ in 2014 [17, 18]. Furthermore, based on the BMI Asian populations classification, in Indonesia, overweight and obesity were more common in women than men. The trend has increased over time, with 61.1% and 42.4% in women and men respectively [17].

To our knowledge, however, there is still a limited understanding of the BMI cut-off point using the Asian population category and stroke and heart disease. Therefore, identifying the association between the BMI categories for Asian population and stroke and heart disease has become an important issue for planning effective strategies to reduce the number of strokes and heart disease among the Indonesian adult population. Thus, the present study aims to predict the incidence rate ratio of stroke and heart disease by BMI categories for Asian population.

## Methods

### Study population

The IFLS is an ongoing longitudinal study conducted in 1993, 1997, 2000, 2007, and 2014. The IFLS survey was conducted using in-person interviews with adult participants. Questionnaires were used to collect information regarding socioeconomics and health [19]. As described in detail elsewhere [19, 20], the survey sample comprised households and represented about 83% of the Indonesian population living in 13 of the 27 total provinces in 1993 [21].

The fourth (2007) and fifth (2014) waves of the Indonesia Family Life Survey (IFLS) were used in this longitudinal study. The IFLS survey 2007 was conducted between November 2007 and mid-July 2008, and the IFLS survey 2014 from September 2014 until April 2015 [19, 20].

In the present study, the eligible population for the 2007 IFLS survey consisted of 14,039 adult participants, both men and women, aged ≥ 35 years, among whom 13,630 responded to the questionnaire in the 2007 IFLS survey. After screening the data under the inclusion and exclusion criteria, the data were examined for duplicate entries and extreme outliers (Figure 1). Among the 2007 IFLS survey participants, we excluded 2,476 with missing data for height and 9 with missing data for weight. We also excluded 95 extreme outliers in height (<100 cm or >200 cm) and weight (<25 kg or >200 kg)[17], 2,273 with missing data for stroke and heart disease, 291 who had stroke and heart disease, and 4 who had duplicate entries.

Next, we excluded 892 who had died before the 2014 IFLS survey, 471 who had moved out of the IFLS areas before the 2014 IFLS survey, and 431 with missing data on stroke and heart disease in the 2014 IFLS survey. Thus, 6,688 participants were analyzed in this study. This was a 7-year longitudinal study between 2007 and 2014. Stroke and heart disease were confirmed for 334 (5.0%) individuals 2014.

### Exposure

The BMI of adults was considered for exposure, which was measured at baseline in 2007. BMI was calculated as body weight (in kilograms) divided by the square of body height (in meters). In the IFLS survey, height and weight were measured by trained interviewers (usually trained nurses) [20]. The Seca Plastic Height Board Model 213 was used to measure heights; the measurement was taken to the nearest millimeter and the Camry model EB1003 scale was used to measure weight to the nearest tenth of a kilogram [19]. The BMI cut-off point of the World Health Organization (WHO) for Asian populations [16] was used; therefore, we classified BMI into four groups: underweight (<18.5 kg/m^2^), or normal weight (18.5–22.9 kg/m^2^), or overweight (23.0– 27.4 kg/m^2^) or obese (≥27.5 kg/m^2^).

### Outcome

Stroke and heart disease were outcome variables that were evaluated both in 2007 and 2014. Stroke and heart disease were assessed with questions, 1) “Has a doctor, paramedic, or nurse ever told you that you had heart disease?”; 2) “Has a doctor, paramedic, or nurse ever told you that you had a stroke?” to which participants answered “yes” or “no.” The stroke and heart disease were then classified as “yes” (i.e., participants who have either stroke or heart disease or both stroke and heart disease) or “no” (i.e., participants who do not have stroke or heart disease or both stroke and heart disease).

### Covariates

Regions, areas, history of diseases (e.g., hypertension, diabetes, lung conditions, and cancer), physical activity, employment status, education level, smoking status, depression status, visit to a health facility, monthly per-capita expenditure (monthly PCE), participation in community activities, blood pressure, C-reactive protein (CRP), high-density lipoprotein cholesterol (HDL-C), and total cholesterol (TC) were treated as covariates, which were assessed at baseline in 2007.

The region was classified into three groups representing province areas in Indonesia (Sumatra, Java-Bali, and West Nusa Tenggara, Central, South and East Kalimantan, South, North, and West Sulawesi). We also categorized areas into two groups, “urban or “rural”, according to the Central Bureau of Statistics [22–24].

Hypertension, diabetes, lung condition, and cancer were assessed with questions, 1) “Has a doctor, paramedic, or nurse ever told you that you had hypertension/diabetes/lung condition/cancer?”classified as yes (i.e., participants who had the disease) or no (i.e., participants who had no disease). Physical activity was measured according to the short version of the International Physical Activity Questionnaire (IPAQ); an explanation of IPAQ has been described in detail elsewhere [25]. We categorized physical activity as moderate-high physical activity or low physical activity.

Employment status was divided as “employed” (i.e., working/helping to earn income) or “unemployed” (combining the categories “job searching”, “housekeeping”, “retired”, and “sick/disabled”). Education level was determined using the following questions: 1) “Have you ever attended school?”, to which participants answered “yes” or “no”; 2) “What is the highest education level attained?”, to which the answer was categorized as elementary, junior high school or equivalent, senior high school or equivalent, or college or university (D1, D2, D3). From 1984 to 1994, all Indonesians were required to complete elementary education [26, 27] (i.e., primary education according to the International Standard Classification of Education (ISCED) 2011 [28]); therefore, we classified education level as below lower-secondary education (no schooling or primary education) or lower-secondary education and above (junior high school or equivalent and above).

Smoking status was measured using the following questions: 1) “Have you ever chewed tobacco, smoked a pipe, smoked self-rolled cigarettes, or smoked cigarettes/cigars?”, to which participants answered “yes” or “no”; 2) “do you still have the habit or have you totally quit?”, to which answered as “still have” or “quit”. We then classified the answer as a smoker, former smoker, or non-smoker.

Depression status was assessed by ten questions from the Center for Epidemiologic Studies Depression (CES-D) scale. The score of the 10 CES-D questionnaire answers with a lowest score of 10 and a highest score of 40. By reason, the score ranged from 10 to 40, the score was rebased to zero to 30, with the highest score designating to the most depression [29]. A prior study suggested the cut-off point for depression was set to a score of ≥10 [29–31]. We then categorized the answers into two groups: not having depression (score <10) or having depression (score ≥10).

Visit to a health facility was divided into two groups: “yes” (i.e., visit to a health facility in the last four weeks) or “no” (i.e., no visit to a health facility in the last four weeks). Health insurance status was categorized as yes (i.e., have health insurance) or no (i.e., do not have health insurance).

Monthly per-capita expenditure (monthly PCE) was calculated as monthly total household expenditure divided by the number of household members according to the definition of the IFLS consumption expenditure [32]. We defined low monthly PCE as the bottom 40% of total monthly PCE according to the definition of poor by the World Bank and WHO [33]. The monthly PCE in the IFLS data was assessed in Indonesian rupiah (IDR), then converted into US dollars (USD) at 2007 exchange rates (1 USD = 9,141 IDR) [34]. Then we categorized monthly PCE into two groups: ≤41 USD (low monthly PCE) or >41 USD (high monthly PCE).

Participation in community activities was considered in two groups: yes (i.e., participants who participated in at least one type of community activity) or no (i.e., participants who never participated in any of the four types of community activities [community meetings, voluntary labor, programs to improve the village/neighborhood, or religious activities]).

Blood pressure (BP) was measured in a seated position using a sphygmomanometer, by regularly trained interviewers. High blood pressure was defined as systolic blood pressure ≥140 mmHg or diastolic ≥90 mmHg. Normal C-reactive protein (CRP) was defined as ≤0.3 mg/dL[35]. Low high-density lipoprotein cholesterol (HDL-C) was defined as serum HDL-C <40 mg/dL in males and <50 mg/dL in females [36]. A borderline high of total cholesterol (TC) was defined as ≥200mg/dL [37].

### Statistical analysis

Descriptive statistics were used to describe the study population’s, study variables and were evaluated using analysis of one-way ANOVA for continuous variables and the chi-square test for categorical variables.

A multivariate-adjusted Poisson regression model was used to estimate the incidence rate ratios (IRR) and 95% confidence intervals (CIs) of either stroke or heart disease or both stroke and heart disease according to BMI. The modeling approach is prediction rather than causal inference. Stroke and heart disease categorized as the dichotomous outcome variable. Multivariate models were adjusted for the following variables. According to the BMI cut-off point for Asian populations [16], BMI was categorized into four groups, and participants with normal weight (BMI 18.0-22.9 kg/m^2^) were defined as a reference category.

Model 1 was adjusted for age (≤44, 45-49, 50-54, 55-59, and ≥60 years) and sex. Model 2 was adjusted for age (≤44, 45-49, 50-54, 55-59, and ≥60 years), sex, regions (Sumatra, Java-Bali, or Sumatra, Java-Bali, and West Nusa Tenggara, Central, South and East Kalimantan, South, North and West Sulawesi), and areas (urban or rural). Model 3 was further adjusted for model 2 plus, diabetes (yes or no), lung condition (yes or no), blood pressure (low or high or missing), employment status (employee or unemployment or missing) and education level (below lower-secondary or lower-secondary education and above or missing). To examine whether the association between BMI and stroke and heart disease was attributable to health behavior, model 4 was further adjusted for model 3, added smoking status (current smoker, or former smoker, or non-smoker or missing), physical activity (high-moderate or low or missing), visit to a health facility (yes or no or missing) and health insurance status (yes or no or missing).

In addition, subgroup analyses were used to predict the incidence rate ratio of stroke and heart disease by BMI categories for Asian population in terms of age (<55 years, and ≥55 years). The age range was based on data from the 2020 Indonesian population census, which indicates that the majority of the labor force is between the ages of 35 and 54 [38].

The Poisson regression model was performed using the Stata Statistical Software: Release 17. College Station, TX: StataCorp LLC. All statistical tests were two-sided, p-values < 0.05 were considered statistically significant.

## Results

**Table 1** shows the baseline characteristics of participants according to BMI categories for Asian population. Participants with obesity (BMI ≥ 27.5 kg/m^2^) were younger and more likely to be female, to live in an urban area, to have hypertension, to have low physical activity, to have a higher education level (lower-secondary education and above), to be non-smokers, and to have higher blood pressure. Participants with obesity are also less likely to not visit a health facility or have health insurance.

**Table 1.**
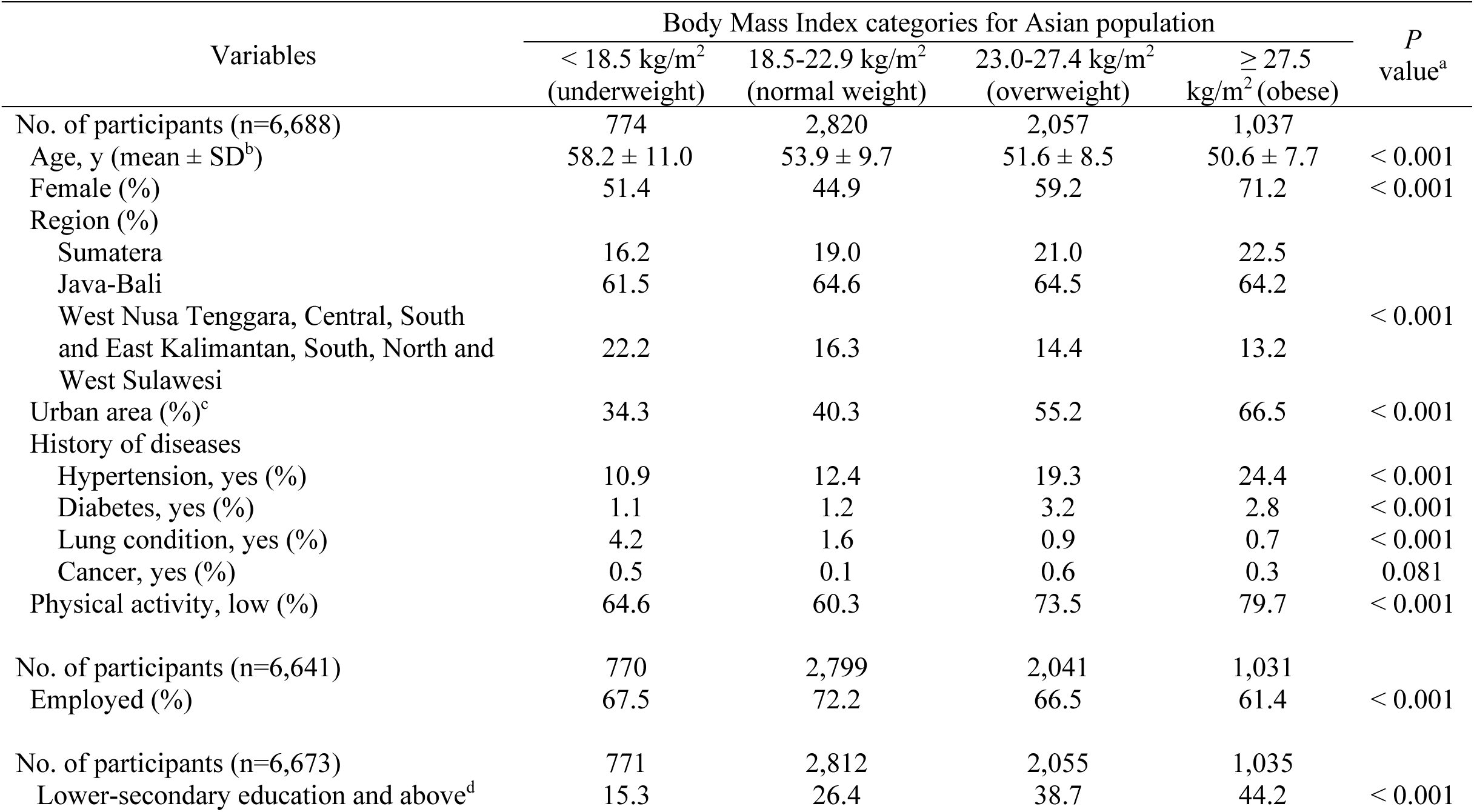
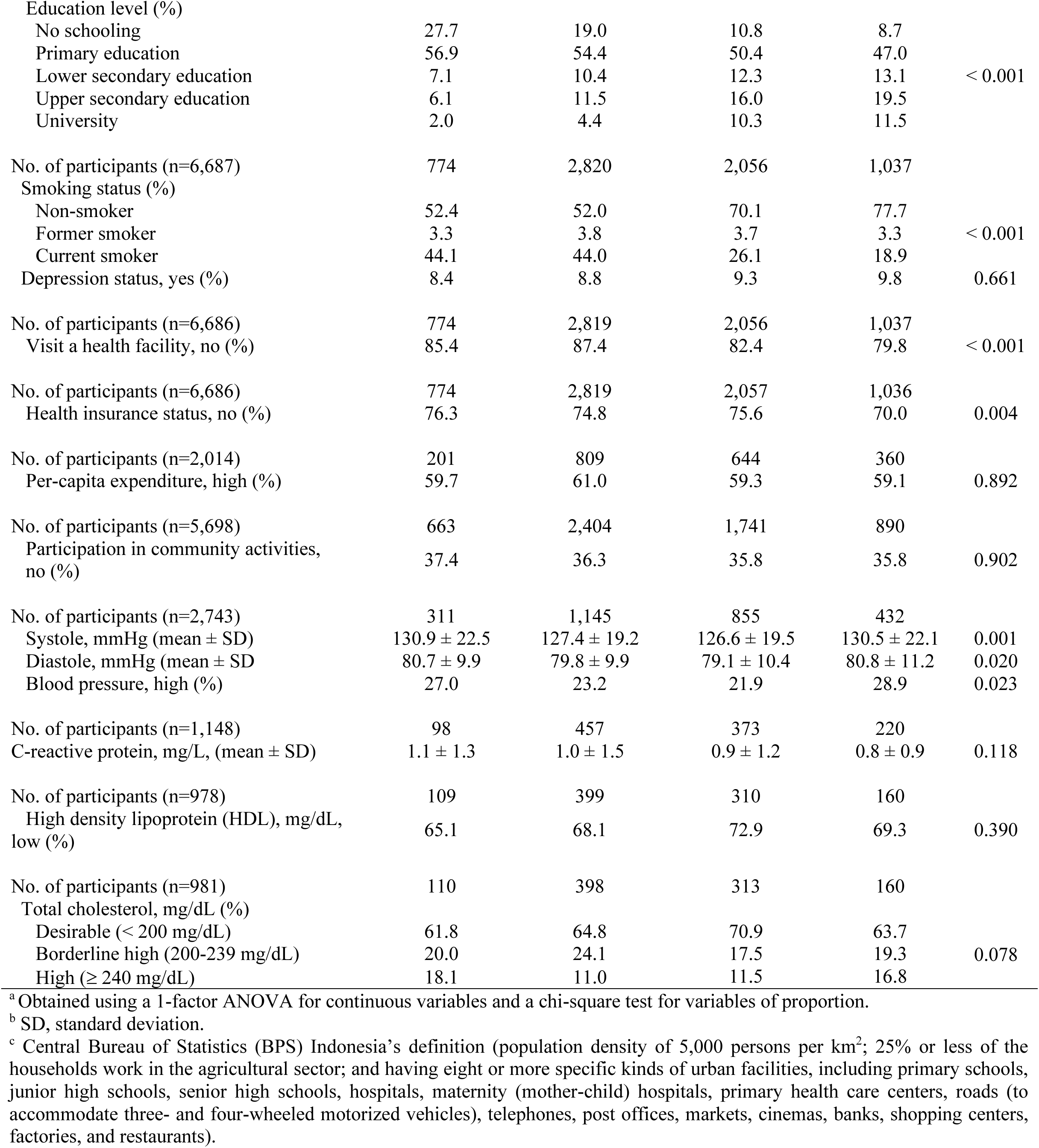
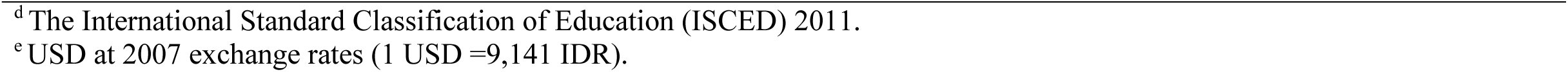
Baseline characteristics of the participants by BMI, IFLS 2007.

| Variables | Body Mass Index categories for Asian population |  |  |  | *P*<br>value<sup>a</sup> |
| --- | --- | --- | --- | --- | --- |
|  | < 18.5 kg/m<sup>2</sup><br>(underweight) | 18.5-22.9 kg/m<sup>2</sup><br>(normal weight) | 23.0-27.4 kg/m<sup>2</sup><br>(overweight) | ≥ 27.5<br>kg/m<sup>2</sup> (obese) |  |
| No. of participants (n=6,688) | 774 | 2,820 | 2,057 | 1,037 |  |
| Age, y (mean ± SD<sup>b</sup>) | 58.2 ± 11.0 | 53.9 ± 9.7 | 51.6 ± 8.5 | 50.6 ± 7.7 | < 0.001 |
| Female (%) | 51.4 | 44.9 | 59.2 | 71.2 | < 0.001 |
| Region (%) |  |  |  |  |  |
| Sumatera | 16.2 | 19.0 | 21.0 | 22.5 |  |
| Java-Bali | 61.5 | 64.6 | 64.5 | 64.2 |  |
| West Nusa Tenggara, Central, South<br>and East Kalimantan, South, North and<br>West Sulawesi | 22.2 | 16.3 | 14.4 | 13.2 | < 0.001 |
| Urban area (%)<sup>c</sup> | 34.3 | 40.3 | 55.2 | 66.5 | < 0.001 |
| History of diseases |  |  |  |  |  |
| Hypertension, yes (%) | 10.9 | 12.4 | 19.3 | 24.4 | < 0.001 |
| Diabetes, yes (%) | 1.1 | 1.2 | 3.2 | 2.8 | < 0.001 |
| Lung condition, yes (%) | 4.2 | 1.6 | 0.9 | 0.7 | < 0.001 |
| Cancer, yes (%) | 0.5 | 0.1 | 0.6 | 0.3 | 0.081 |
| Physical activity, low (%) | 64.6 | 60.3 | 73.5 | 79.7 | < 0.001 |
| No. of participants (n=6,641) | 770 | 2,799 | 2,041 | 1,031 |  |
| Employed (%) | 67.5 | 72.2 | 66.5 | 61.4 | < 0.001 |
| No. of participants (n=6,673) | 771 | 2,812 | 2,055 | 1,035 |  |
| Lower-secondary education and above<sup>d</sup> | 15.3 | 26.4 | 38.7 | 44.2 | < 0.001 |
| Education level (%) |  |  |  |  |  |
| No schooling | 27.7 | 19.0 | 10.8 | 8.7 |  |
| Primary education | 56.9 | 54.4 | 50.4 | 47.0 |  |
| Lower secondary education | 7.1 | 10.4 | 12.3 | 13.1 | < 0.001 |
| Upper secondary education | 6.1 | 11.5 | 16.0 | 19.5 |  |
| University | 2.0 | 4.4 | 10.3 | 11.5 |  |
| No. of participants (n=6,687) | 774 | 2,820 | 2,056 | 1,037 |  |
| Smoking status (%) |  |  |  |  |  |
| Non-smoker | 52.4 | 52.0 | 70.1 | 77.7 |  |
| Former smoker | 3.3 | 3.8 | 3.7 | 3.3 | < 0.001 |
| Current smoker | 44.1 | 44.0 | 26.1 | 18.9 |  |
| Depression status, yes (%) | 8.4 | 8.8 | 9.3 | 9.8 | 0.661 |
| No. of participants (n=6,686) | 774 | 2,819 | 2,056 | 1,037 |  |
| Visit a health facility, no (%) | 85.4 | 87.4 | 82.4 | 79.8 | < 0.001 |
| No. of participants (n=6,686) | 774 | 2,819 | 2,057 | 1,036 |  |
| Health insurance status, no (%) | 76.3 | 74.8 | 75.6 | 70.0 | 0.004 |
| No. of participants (n=2,014) | 201 | 809 | 644 | 360 |  |
| Per-capita expenditure, high (%) | 59.7 | 61.0 | 59.3 | 59.1 | 0.892 |
| No. of participants (n=5,698) | 663 | 2,404 | 1,741 | 890 |  |
| Participation in community activities, no (%) | 37.4 | 36.3 | 35.8 | 35.8 | 0.902 |
| No. of participants (n=2,743) | 311 | 1,145 | 855 | 432 |  |
| Systole, mmHg (mean ± SD) | 130.9 ± 22.5 | 127.4 ± 19.2 | 126.6 ± 19.5 | 130.5 ± 22.1 | 0.001 |
| Diastole, mmHg (mean ± SD) | 80.7 ± 9.9 | 79.8 ± 9.9 | 79.1 ± 10.4 | 80.8 ± 11.2 | 0.020 |
| Blood pressure, high (%) | 27.0 | 23.2 | 21.9 | 28.9 | 0.023 |
| No. of participants (n=1,148) | 98 | 457 | 373 | 220 |  |
| C-reactive protein, mg/L, (mean ± SD) | 1.1 ± 1.3 | 1.0 ± 1.5 | 0.9 ± 1.2 | 0.8 ± 0.9 | 0.118 |
| No. of participants (n=978) | 109 | 399 | 310 | 160 |  |
| High density lipoprotein (HDL), mg/dL, low (%) | 65.1 | 68.1 | 72.9 | 69.3 | 0.390 |
| No. of participants (n=981) | 110 | 398 | 313 | 160 |  |
| Total cholesterol, mg/dL (%) |  |  |  |  |  |
| Desirable (< 200 mg/dL) | 61.8 | 64.8 | 70.9 | 63.7 |  |
| Borderline high (200-239 mg/dL) | 20.0 | 24.1 | 17.5 | 19.3 | 0.078 |
| High (≥ 240 mg/dL) | 18.1 | 11.0 | 11.5 | 16.8 | |

<sup>a</sup> Obtained using a 1-factor ANOVA for continuous variables and a chi-square test for variables of proportion.
<sup>b</sup> SD, standard deviation.
<sup>c</sup> Central Bureau of Statistics (BPS) Indonesia's definition (population density of 5,000 persons per km<sup>2</sup>; 25% or less of the households work in the agricultural sector; and having eight or more specific kinds of urban facilities, including primary schools, junior high schools, senior high schools, hospitals, maternity (mother-child) hospitals, primary health care centers, roads (to accommodate three- and four-wheeled motorized vehicles), telephones, post offices, markets, cinemas, banks, shopping centers, factories, and restaurants).
<sup>d</sup> The International Standard Classification of Education (ISCED) 2011.
<sup>e</sup> USD at 2007 exchange rates (1 USD =9,141 IDR).

The incidence rate ratio of stroke and heart disease by BMI is shown in **Table 2**. We found that the incidence rate ratio of stroke and heart disease was significantly higher among participants with obesity (BMI ≥ 27.5 kg/m^2^) as compared to participants with normal weight (18.0-22.9 kg/m^2^) in model 1 (P<0.001). Even with the addition of several adjustment items, these associations remained significant. The multivariate-adjusted incidence rate ratio (IRR) (95% confidence interval (CI)) was 2.57 (1.64-4.04) (Model 4; P<0.001).

**Table 2.**
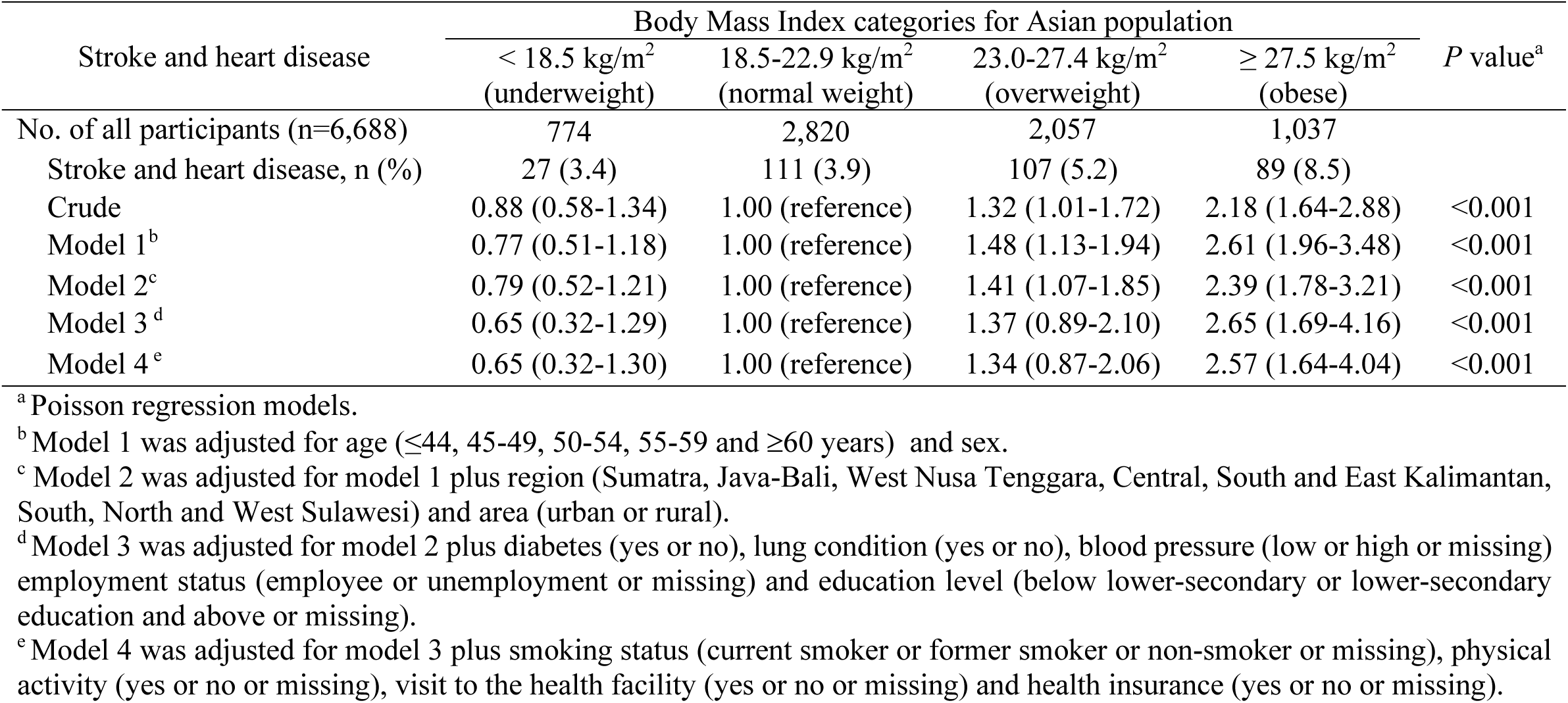
Incidence rate ratio of stroke and heart disease by BMI, IFLS 2007 and 2014.

**Table 3** shows the incidence rate ratio of stroke and heart disease by BMI, stratified by age. We found that for age <55 years and BMI ≥ 27.5 kg/m^2^, the percentage of stroke and heart disease is 7.2%, which is lower than for those with age >=55 years and BMI ≥ 27.5 kg/m^2^ (12.3%) however, their incidence rate ratio is higher (4.18 vs. 1.53).

**Table 3.** Incidence rate ratio of stroke and heart disease by BMI, stratified by age, IFLS 2007 and 2014.

|  | Body Mass Index categories for Asian population |  |  |  | P value<sup>a</sup> |
| --- | --- | --- | --- | --- | --- |
|  | < 18.5 kg/m<sup>2</sup><br>(underweight) | 18.5-22.9 kg/m<sup>2</sup><br>(normal weight) | 23.0-27.4 kg/m<sup>2</sup><br>(overweight) | ≥ 27.5 kg/m<sup>2</sup><br>(obese) |  |
| Age <55 years (n=4,277) |  |  |  |  |  |
| No of participants | 340 | 1,707 | 1,460 | 770 |  |
| Stroke and heart disease, n (%) | 5 (1.4) | 44 (2.5) | 67 (4.5) | 56 (7.2) |  |
| Crude | 0.57 (0.22-1.43) | 1.00 (reference) | 1.78 (1.21-2.60) | 2.82 (1.90-4.18) | <0.001 |
| Model 1<sup>b</sup> | 0.57 (0.22-1.43) | 1.00 (reference) | 1.78 (1.21-2.61) | 2.82 (1.88-4.23) | <0.001 |
| Model 2<sup>c</sup> | 0.58 (0.23-1.46) | 1.00 (reference) | 1.70 (1.16-2.50) | 2.59 (1.72-3.92) | <0.001 |
| Model 3<sup>d</sup> | 0.78 (1.77-3.45) | 1.00 (reference) | 1.82 (0.92-3.58) | 4.25 (2.14-8.43) | <0.001 |
| Model 4<sup>e</sup> | 0.81 (0.18-3.69) | 1.00 (reference) | 1.75 (0.88-3.47) | 4.18 (2.10-8.31) | 0.001 |
| Age ≥55 years (n=2,411) |  |  |  |  |  |
| No of participants | 434 | 1,113 | 597 | 267 |  |
| Stroke and heart disease, n (%) | 22 (5.0) | 67 (6.0) | 40 (6.7) | 33 (12.3) |  |
| Crude | 0.84 (0.52-1.36) | 1.00 (reference) | 1.11 (0.75-1.64) | 2.05 (1.35-3.11) | 0.004 |
| Model 1<sup>b</sup> | 0.85 (0.53-1.39) | 1.00 (reference) | 1.16 (0.78-1.72) | 2.21 (1.45-3.37) | 0.001 |
| Model 2<sup>c</sup> | 0.88 (0.54-1.42) | 1.00 (reference) | 1.09 (0.73-1.63) | 2.02 (1.31-3.12) | 0.001 |
| Model 3<sup>d</sup> | 0.61 (0.27-1.33) | 1.00 (reference) | 1.10 (0.62-1.95) | 1.55 (0.79-3.02) | 0.116 |
| Model 4<sup>e</sup> | 0.61 (0.28-1.35) | 1.00 (reference) | 1.12 (0.63-1.98) | 1.53 (0.78-3.02) | 0.129 |

<sup>a</sup> Poisson regression models.
<sup>b</sup> Model 1 was adjusted for sex.
<sup>c</sup> Model 2 was adjusted for model 1 plus region (Sumatra, Java-Bali, West Nusa Tenggara, Central, South and East Kalimantan, South, North and West Sulawesi) and area (urban or rural).
<sup>d</sup> Model 3 was adjusted for model 2 plus diabetes (yes or no), lung condition (yes or no), blood pressure (low, high or missing) employment status (employee or unemployment or missing) and education level (below lower-secondary or lower-secondary education and above or missing).
<sup>e</sup> Model 4 was adjusted for model 3 plus smoking status (current smoker or former smoker or non-smoker or missing), physical activity (yes or no or missing), visit to the health facility (yes or no or missing) and health insurance (yes or no or missing).

## Discussion

In this longitudinal study, we investigated the incidence rate ratio of stroke and heart disease by BMI categories for Asian population. We found a significant incidence rate ratio of stroke and heart disease by BMI categories. To our knowledge, the present study is one of the first studies to investigate the incidence rate ratio of stroke and heart disease by BMI categories for Asian population among adult Indonesian Individuals.

Previous studies have examined the association between BMI and outcomes directly related to our definition of stroke and heart disease. However, generally, BMI was classified in the earlier study using WHO cut-off points. As in a prior study in Japan, where BMI was divided into seven categories: <19, 19–<21, 21–<23, 23–<25, 25–<27, 27–<30, ≥30 kg/m^2^, it was reported that a higher cumulative average BMI was associated with ischemic stroke both in men and women [11]. Also, in a previous study in China, higher BMI (<18.0, 18.0-20.4, 20.5-22.9, 23.0-24.9, 25.0-27.4, 27.5-29.9, and >30.0 kg/m^2^) was positively associated with ischemic stroke [39]. Another study in China also indicated that being overweight (24–27.9 kg/m^2^) at baseline increased the risk of both ischemic and hemorrhagic strokes; obesity (≥28 kg/m^2^) increased the risk of ischemic stroke [40]. Regarding the association between BMI and heart disease, previous European studies reported that a higher BMI causes an increased risk of an individual having heart disease [41], and increasing the severity of obesity would increase CVDs and mortality [42]. Also, having a stable overweight or obesity throughout adulthood was associated with increased chronic heart disease (CHD) risk [43]. Another study in Australia also found that adults with overweight condition or obesity increased their risk of CVDs [44]. This was in line with our findings when we used BMI for the Asian population cut-off point, where the obesity cut-off points were lower than BMI WHO cut-off points. Thus, The BMI Asian population cut-off point might be appropriate to use to find the association between BMI and health outcomes in Asians as well as Indonesians.

We also investigated the incidence rate ratio of stroke and heart disease by BMI categories for Asian population stratified by age. We found that the incidence rate ratio of stroke and heart disease was higher in middle-aged (age <55 years old); however, the incidence rate ratio was attenuated in the older category (age ≥55 years old). This discrepancy could be attributed to disparities in socioeconomic status (education level and employment) [6], unhealthy lifestyle behaviour (i.e., unhealthy diet and low physical activity) [45], and living area among age groups [18]. Therefore, we performed a living area-stratified analysis among age groups in order to examine the possibility that differences in living area between middle- and older-aged participants might affect the incidence rate ratio of stroke and heart disease. We observed that obese middle-aged (age<55 years old) who live in urban or rural areas have a higher incidence rate ratio of stroke and heart disease compared with middle-age with normal weight (see supplementary Table S1). However, the results among older (age ≥55 years old) show a significant change only for those living in rural areas (see supplementary Table S2).

Over fifty percent of Indonesians currently reside in urban areas [18]. Furthermore, according to the 2020 Population Census, 79.7% of the workforce is between the ages of 35 and 54 [38]. Education, on the other hand, may have a higher impact on the employment of the middle-aged since they are more likely to work in skilled labor positions [18, 46]. In contrast, older individuals are more likely to work in agriculture [47]. In line with our study, when further looking at our baseline characteristics by age, we found that participants with ages <55 years were more likely to be employees, have a higher education level (lower-secondary education and above), and live in an urban area.

In addition, a previous study in Indonesia suggested that higher education, and white scholar workers were associated with higher BMI [18]. The reasons for this might be in Indonesia, higher education is related to better socioeconomic status [17]; however, socioeconomic level was not significantly influenced by self-reported health status [48], and people with higher socioeconomic status are more likely to experience health issues like obesity and hypertension [49, 50]. Furthermore, greater BMI in the middle age category has been linked to an increase in stroke and myocardial infraction [51], the highest association with incident heart failure among CVD subtypes [52], and may increase the risk of heart disease [41], which is in line with our findings. Therefore, it is important to encourage middle-aged individuals to reduce BMI as a preventive against non-communicable diseases, especially stroke and heart disease.

The strengths of the present study are as follows. First, this was one of the first known longitudinal study to investigate the incidence rate ratio of stroke and heart disease by BMI categories for Asian population. Second, since this study used BMI categories for Asian population, the findings can be more sensitive to capture over and undernutrition among populations in Asian countries. Third, the IFLS used calibrated tools and trained interviewers to measure height and weight.

However, this study also has several limitations. First, we used self-reported data for stroke and heart disease, which may cause information bias. Second, data on BMI were only examined at baseline in 2007, and information on BMI may have changed between the 2007 and the 2014 IFLS survey. Third, even though this study represents the Indonesian population, this result might be underestimated because the analysis was taken from all participants who survived diseases. Fourth, this study might not be representative of the Asian population because the participants were solely drawn from Indonesian individuals.

## Conclusions

Our results indicate the BMI Asian population cut-off point is suitable to determine the association between BMI and health outcomes in Indonesia and also in other Asian nations that have similar characteristics to Indonesia. Additionally, according to our findings, the incidence rate ratio of stroke and heart disease was higher in middle-aged obese individuals rather than in older obese individuals. Our study suggests that lowering BMI through healthy food and increasing physical exercise among middle-aged individuals are needed to prevent stroke and heart disease, especially for those with high education, who are employees, and who live in urban or rural areas.

## Data Availability

All data produced are available online at

https://www.rand.org/well-being/social-and-behavioral-policy/data/FLS/IFLS/access.html

## Acknowledgments

This work was supported by Taipei Medical University. We also appreciate the RAND Corporation for making the data accessible to the general public.

## Authors’ contribution

Study concept and design: KL, WG, DN; Analyzed the data: KL, DN, MKT; Drafting of the manuscript: KL; Interpretation of the data and revision of the manuscript: KL, WG, DN, MKT; All authors read and approved the final manuscript.

## Data availability

Data is publicly available at https://www.rand.org/well-being/social-and-behavioral-policy/data/FLS/IFLS/access.html.

## Declarations

### Ethics approval and consent to participate

The IFLS data used in the present study are publicly available. The surveys and procedures of the IFLS study were reviewed and approved by institutional review boards (IRBs) at the RAND Corporation in the US and the University of Gadjah Mada (UGM) in Indonesia with the ethical clearance No. s0064-06-01-CR01. Written informed consent was obtained from all participants before data collection began.

### Consent for publication

None.

### Competing interest

The authors declare no conflict of interest.

### Funding

None.

## List of abbreviation

BMI: Body mass index
IFLS: Indonesia Family Life Survey
GBD: Global Burden of Disease
IRR: Incidence rate ratio
CIs: Confidence intervals
CVDs: Cardiovascular diseases
IHD: Ischemic heart disease
DALYS: Death and disability
NCDs: Non-communicable diseases
WHO: World Health Organization
IPAQ: International Physical Activity Questionnaire
ISCED: International Standard Classification of Education
CES-D: Center for Epidemiologic Studies Depression
PCE: Per-capita expenditure
IDR: Indonesian rupiah
USD: US dollars
CRP: C-reactive protein
HDL-C: High-density lipoprotein cholesterol
TC: Total cholesterol
CHD: Chronic heart disease

## Notes

### Competing Interest Statement

The authors have declared no competing interest.

### Funding Statement

This study did not receive any funding

